# Estimating unobserved SARS-CoV-2 infections in the United States

**DOI:** 10.1101/2020.03.15.20036582

**Authors:** T. Alex Perkins, Sean M. Cavany, Sean M. Moore, Rachel J. Oidtman, Anita Lerch, Marya Poterek

## Abstract

By March 2020, COVID-19 led to thousands of deaths and disrupted economic activity worldwide. As a result of narrow case definitions and limited capacity for testing, the number of unobserved SARS-CoV-2 infections during its initial invasion of the US remains unknown. We developed an approach for estimating the number of unobserved infections based on data that are commonly available shortly after the emergence of a new infectious disease. The logic of our approach is, in essence, that there are bounds on the amount of exponential growth of new infections that can occur during the first few weeks after imported cases start appearing. Applying that logic to data on imported cases and local deaths in the US through March 12, we estimated that 22,876 (95% posterior predictive interval: 7,451 - 53,044) infections occurred in the US by this date. By comparing the model’s predictions of symptomatic infections to local cases reported over time, we obtained daily estimates of the proportion of symptomatic infections detected by surveillance. This revealed that detection of symptomatic infections decreased throughout February as exponential growth of infections outpaced increases in testing. Between February 21 and March 12, we estimated an increase in detection of symptomatic infections, which was strongly correlated (median: 0.97, 95% PPI: 0.85 - 0.98) with increases in testing. These results suggest that testing was a major limiting factor in assessing the extent of SARS-CoV-2 transmission during its initial invasion of the US.

**Significance Statement:** Countries across the world observed dramatic rises in COVID-19 cases and deaths in March 2020. In the United States, delays in the availability of diagnostic testing prompted questions about the extent of unobserved community transmission. Using a simulation model informed by reported cases and deaths, we estimated that tens of thousands of people were infected by the time a national emergency was declared on March 13. Our results indicate that fewer than 20% of locally acquired, symptomatic infections in the US were detected over a period of a month. The existence of a large, unobserved reservoir of infection argues for the necessity of large-scale social distancing that went into effect to mitigate the impacts of SARS-CoV-2 on the US.

## Main Text

## Introduction

SARS-CoV-2 is a newly emerged coronavirus that is causing a global pandemic (1). The unprecedented spread of SARS-CoV-2 owes to its high transmissibility (2), pre-symptomatic transmission (3), and transmission by asymptomatic infections (4). An appreciable fraction of infections are asymptomatic (5), and many others result in mild symptoms that could be mistaken for other respiratory illnesses (6). These factors point to a potentially large reservoir of unobserved infections (7), especially in settings where capacity to test for SARS-CoV-2 has been limited (8). The United States is one such country in which limited testing has been a major concern, particularly as imported cases, and now local cases, have increased over time (9). Until February 27, testing criteria in the US were limited to close contacts of confirmed cases and those with recent travel to China (9). This means that any local infections resulting from an unobserved imported infection would have gone unnoticed. Community transmission occurred without notice while testing was still being rolled out (10, 11), albeit to an unknown extent.

Our goal was to estimate the extent of community transmission of SARS-CoV-2 in the US that occurred prior to its widespread recognition. Unlike other countries where testing and containment measures were pursued aggressively (12, 13), rollout of testing in the US was slow (9) and widespread social-distancing measures did not go into effect until several weeks after the first reported case (14, 15). Understanding the extent of community transmission has major implications for the effectiveness of different options for control (16) and for anticipating the trajectory and impact of the pandemic (17).

## Results

To estimate the extent of community transmission of SARS-CoV-2 in the US, we used a stochastic simulation model that combined importation and local transmission processes. We informed model parameters with estimates from other countries, where available (Table 1), and estimated values of two unknown parameters by fitting the model to data on local reported deaths in the US (18). To model importation, we simulated observed and unobserved imported infections based on the number and timing of imported cases reported in the US (19) and assumptions about the proportion of different infection outcomes (5, 20). To model local transmission, we used a branching process model informed by estimates of the serial interval and reproduction number of SARS-CoV-2 from Singapore (3). Due to aggressive containment efforts there (12), we considered our model to be a conservative representation of community transmission in the US. To relate our model’s predictions to US data on reported cases and deaths, we also simulated the timing of symptom onset (3), case reporting (18), and death (21), for simulated infections for which those outcomes occurred.

**Table 1.**
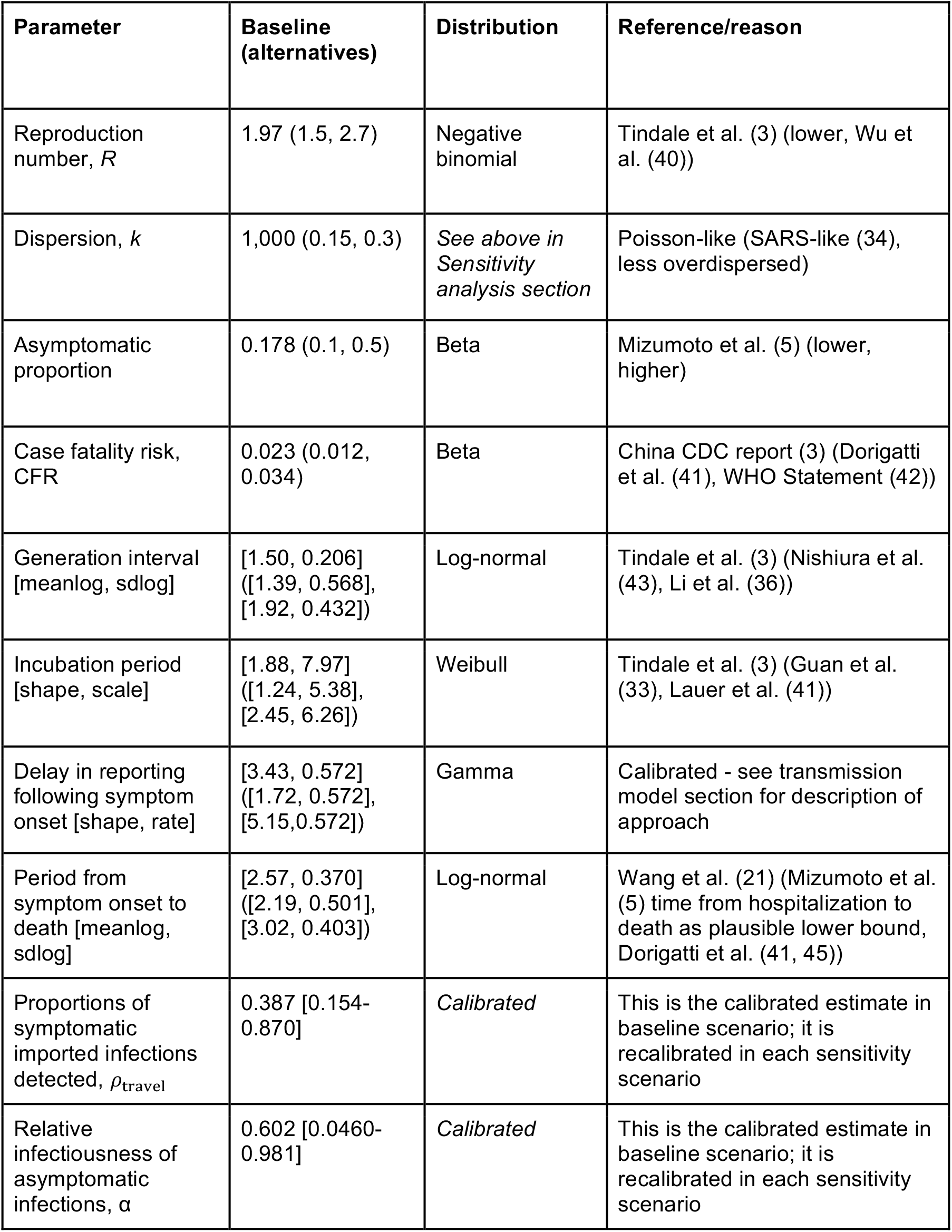
Model parameters. All time periods are given in days.

| Parameter | Baseline (alternatives) | Distribution | Reference/reason |
| --- | --- | --- | --- |
| Reproduction number, $R$ | 1.97 (1.5, 2.7) | Negative binomial | Tindale et al. (3) (lower, Wu et al. (40)) |
| Dispersion, $k$ | 1,000 (0.15, 0.3) | *See above in Sensitivity analysis section* | Poisson-like (SARS-like (34), less overdispersed) |
| Asymptomatic proportion | 0.178 (0.1, 0.5) | Beta | Mizumoto et al. (5) (lower, higher) |
| Case fatality risk, CFR | 0.023 (0.012, 0.034) | Beta | China CDC report (3) (Dorigatti et al. (41), WHO Statement (42)) |
| Generation interval [meanlog, sdlog] | [1.50, 0.206]<br>([1.39, 0.568],<br>[1.92, 0.432]) | Log-normal | Tindale et al. (3) (Nishiura et al. (43), Li et al. (36)) |
| Incubation period [shape, scale] | [1.88, 7.97]<br>([1.24, 5.38],<br>[2.45, 6.26]) | Weibull | Tindale et al. (3) (Guan et al. (33), Lauer et al. (41)) |
| Delay in reporting following symptom onset [shape, rate] | [3.43, 0.572]<br>([1.72, 0.572],<br>[5.15, 0.572]) | Gamma | Calibrated - see transmission model section for description of approach |
| Period from symptom onset to death [meanlog, sdlog] | [2.57, 0.370]<br>([2.19, 0.501],<br>[3.02, 0.403]) | Log-normal | Wang et al. (21) (Mizumoto et al. (5) time from hospitalization to death as plausible lower bound, Dorigatti et al. (41, 45)) |
| Proportions of symptomatic imported infections detected, $\rho_{\text{travel}}$ | 0.387 [0.154-0.870] | *Calibrated* | This is the calibrated estimate in baseline scenario; it is recalibrated in each sensitivity scenario |
| Relative infectiousness of asymptomatic infections, $\alpha$ | 0.602 [0.0460-0.981] | *Calibrated* | This is the calibrated estimate in baseline scenario; it is recalibrated in each sensitivity scenario |

By March 12, there were a total of 1,514 reported cases and 39 reported deaths that resulted from local transmission of SARS-CoV-2 in the US. We used this information to estimate the probability of detecting imported symptomatic infections, ρ_travel_, by seeding our model with imported infections, simulating local transmission, and comparing simulated and reported local deaths. Under our baseline scenario, this resulted in a median estimate of ρ_travel_ = 0.39 (95% posterior predictive interval: 0.15 - 0.90). Simulating from January 1, we obtained 22,876 (95% PPI: 7,451 - 53,044) local infections cumulatively in the US by March 12 (Fig. 1A). Due to the exponential growth posited by our model, 2,958 (95% PPI: 956 - 7,249) local infections were predicted to have occurred on March 12 alone (Fig. 1B). Had we performed a simple extrapolation of reported cases and deaths based on ρ_travel_, our estimate of cumulative local infections by March 12 would have been only 5,018 (95% PPI: 2,350 - 12,445). This suggests that detection of local infections was less sensitive than detection of imported infections.

**Figure 1.**
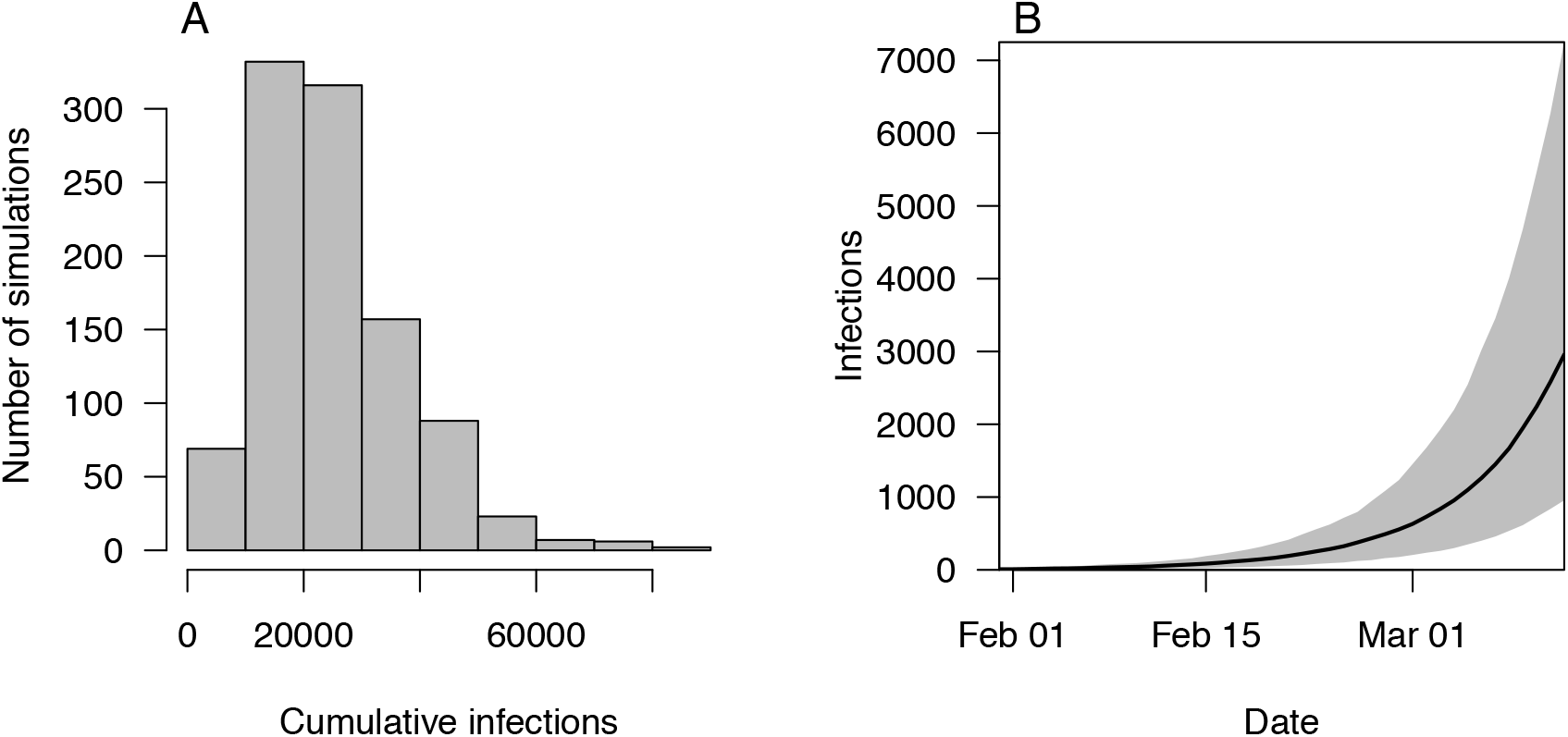
Local infections of SARS-CoV-2 in the US by March 12. These results derive from our baseline scenario and show A) cumulative and B) daily incidence of all local infections, including observed and unobserved. In B, black shows the median and gray shading shows the 95% PPI.

We estimated the probability of detecting local symptomatic infections, ρ_local_, by comparing our model’s predictions of symptomatic infections to local case reports on a daily basis. Over the course of February, daily estimates of ρ_local_ decreased from our uniform prior down to a low of 0.033 (95% PPI: 0.012 - 0.12) on February 21, as increases in simulated local infections outpaced newly reported local cases (Fig. 2A). Our results indicate that detection of symptomatic infections was below 20% for nearly a month (median: 29 days, 95% PPI: 17 - 37 days) when containment still might have been feasible. As testing increased in March (Fig. 2B, red), so too did reported cases (Fig. 2A, red) and daily estimates of ρ_local_ (Fig. 2B, black). By March 12, we estimated ρ_local_ to be 0.77 (95% PPI: 0.29 - 1.00). Between February 21 (low estimate of ρ_local_) and March 12, our daily estimates of ρ_local_ were well correlated with daily numbers of tests administered (Pearson’s correlation, median: 0.97, 95% PPI: 0.85 - 0.98).

**Figure 2.**
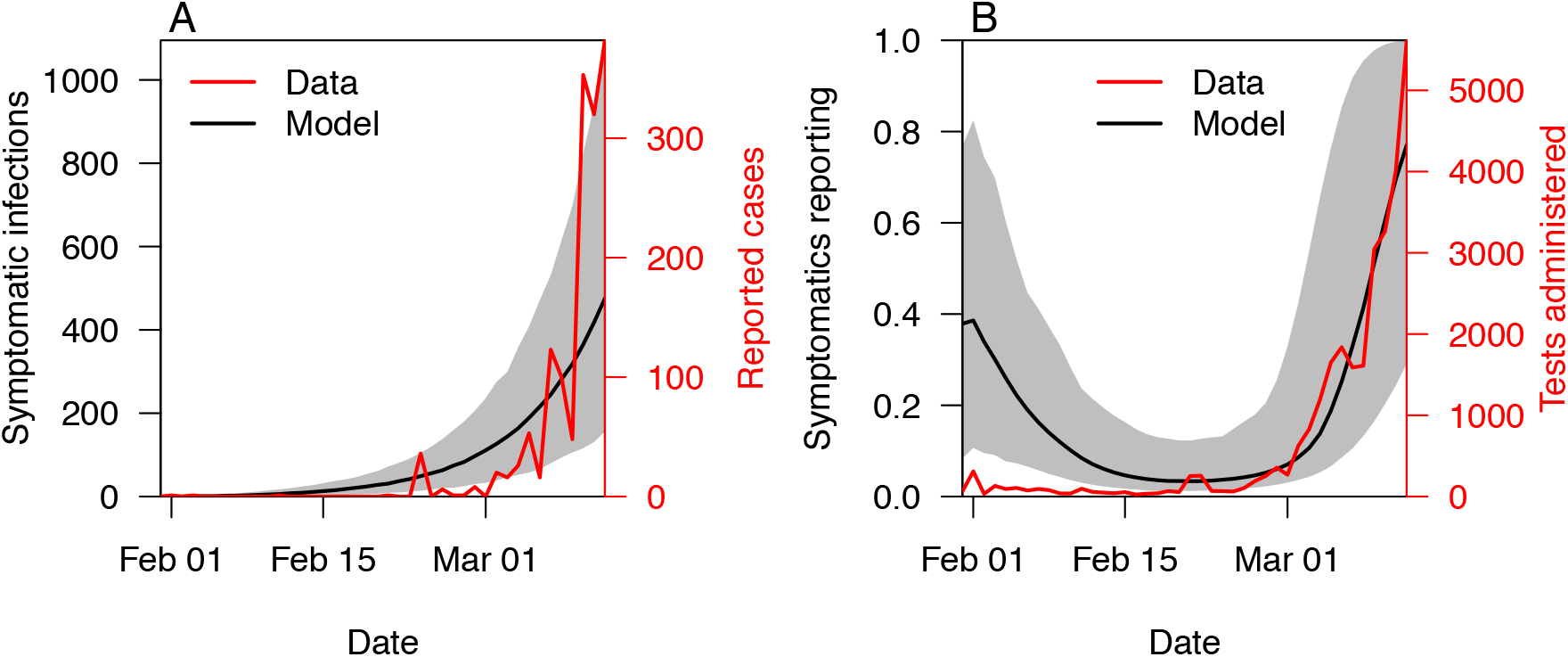
Comparison of symptomatic infections and reported cases. A) Local symptomatic infections predicted under the baseline scenario increased exponentially, whereas reported cases increased more sharply in March. B) Based on this, we estimated how the probability of detecting local symptomatic infections changed daily in the US. Black lines show the median and gray shading shows the 95% PPI.

Successful fitting of our model was demonstrated by its predictions of local deaths by March 12 (median: 33, 95% PPI: 9 - 74), which were consistent with the 39 reported (Fig. 3). Although we did not fit our model to deaths on a daily basis, 85.5% of the deaths predicted by our model occurred within the same range of days over which local deaths were reported (February 29 - March 12). This indicates that, collectively, our model’s assumptions about the timing of importation, local transmission, and delay between exposure and death are plausible. Deaths caused by COVID-19 often occur several weeks after exposure (22). Thus, our baseline model predicted that there would be a median of 395 (95% PPI: 125 - 948) additional deaths as a result of infections that occurred by March 12. Relative to deaths reported by then, this represents an increase by a factor of 12.2 (95% PPI: 7.03 - 21.3).

**Figure 3.**
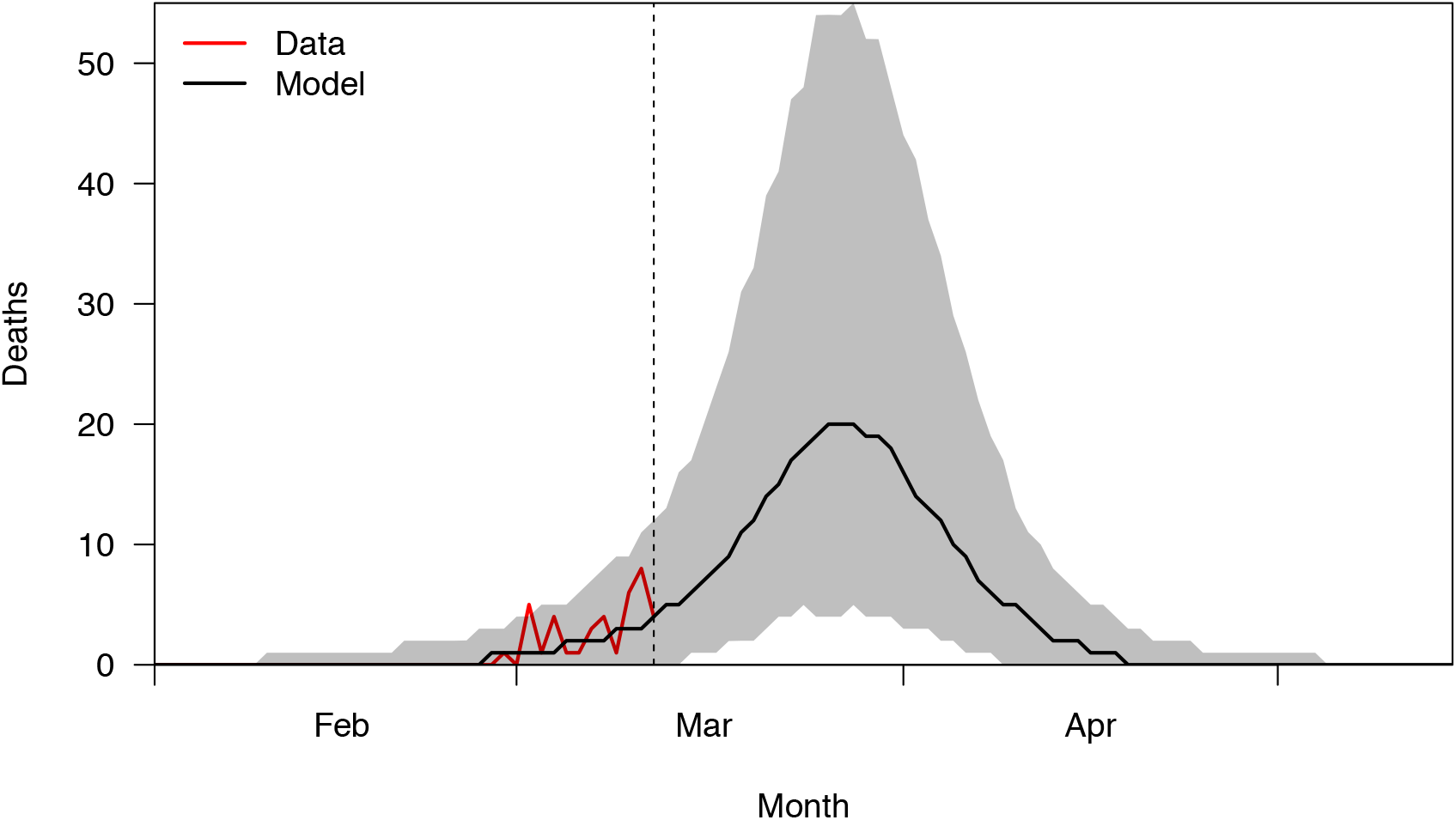
Deaths over time. Our baseline model’s predictions were A) consistent with reported deaths through March 12 (dashed line) and B) indicate that many more deaths should be expected after then based solely on infections that occurred by March 12. Results to the right of the dashed line do not reflect additional deaths that would result from new infections occurring March 13 or after. The black line shows the median and gray shading shows the 95% PPI.

## Discussion

Our approach used a mathematical model to leverage available data to answer a question of significant interest to public health during the initial phase of the COVID-19 pandemic in the US. The only requirements for applying this approach are basic epidemiological data and estimates of standard epidemiological parameters, both of which are collected routinely during the initial weeks of nascent epidemics. Although other approaches – namely, serological surveys – could have provided more direct answers to the question of how many unobserved infections there were in the weeks following the arrival of SARS-CoV-2 in the US, serological assays were only beginning to be developed at that time (23). Relative to other approaches, ours offers the ability to quickly obtain provisional estimates of the number of unobserved infections early in an epidemic, when there still might be time to act on that information with testing and case isolation.

Despite the advantages of our approach, there are limitations of it that should be acknowledged. First, our results were, in some cases, sensitive to deviations from baseline assumptions (Supplementary Information Text). Although most parameter scenarios we explored resulted in similar cumulative infections, higher values of R and earlier importation resulted in estimates in excess of 100,000 (Fig. S6). Second, our branching process model assumes exponential growth, which could be affected by social distancing (24) or the buildup of immunity (25). Neither of those factors were likely to have had much influence on local transmission of SARS-CoV-2 in the US before March 13, however. Third, our parameter assumptions were based on analyses of data collected outside the US. Similar information has proven useful for other pathogens though, such as Zika and Ebola in past public health emergencies (26, 27). Fourth, we did not make use of airline data to model importation (28), but future applications of our method could incorporate that type of information.

The limitations of our approach mean that results from our baseline scenario should be interpreted cautiously. Nonetheless, based on our sensitivity analysis, we conclude that unobserved SARS-CoV-2 infections in the US by March 12 likely numbered in the tens of thousands, and quite possibly in excess of 100,000. This result, considered together with extensive pre-symptomatic and asymptomatic transmission of SARS-CoV-2 (3, 4), suggests that the US was well past the possibility of containment by March 12. Other modeling work (16) suggests that the feasibility of containing SARS-CoV-2 is highly sensitive to the number of infections that occur prior to initiation of containment efforts. Our estimate that fewer than 20% of local symptomatic infections were detected by surveillance for much of February suggests that a crucial opportunity to limit the impact of SARS-CoV-2 on the US may have been missed. Although the number of tests administered increased in March (9), so too did the number of infections and, consequently, the demand for testing.

Coincident with the March 13 declaration of a national emergency (14), social-distancing measures went into effect across the US (15). Our estimate of several thousand active SARS-CoV-2 infections at that time suggests that large-scale mitigation efforts, rather than reactionary measures (29), were indeed necessary. Even after those efforts eventually begin to reverse increases in SARS-CoV-2 transmission in the US, our results show that a downturn in COVID-19 deaths would not be expected to appear until several weeks later. Analyses of the impact of large-scale mitigation efforts in China (30, 31) provide reason for optimism that those measures can be effective.

## Materials and Methods

We calibrated a stochastic model, including separate importation and local transmission steps, to two publicly available datasets on cases of COVID-19 internationally and in the United States. All code and data used are available at http://github.com/TAlexPerkins/sarscov2_unobserved.

### Data

We obtained data on the number of imported cases and deaths from line list data compiled by the Models of Infectious Disease Agent Spread (MIDAS) Network (19). These data informed the number and timing of imported infections predicted by our importation model. We obtained data on the total number of US cases and deaths and total number of cases and deaths globally from time series compiled by the Johns Hopkins University Center for Systems Science and Engineering (18). These data informed our estimates of the proportion of local infections detected. We also used these data in an alternative importation scenario in which the timing of imported infections was sampled proportional to daily global incidence.

### Importation model

We considered cases associated with international travel in the MIDAS dataset to be imported. We removed SARS-CoV-2-positive individuals who were repatriated from the Diamond Princess cruise ship from our analysis, due to the fact that they were quarantined (32), leaving 153 imported cases (including one death). We first estimated the number of imported infections based on the probability that an infection would be symptomatic, the probability of an imported symptomatic infection being detected, and the probability of death among symptomatic infections (case fatality risk, CFR). The CFR and the probability that an infection is symptomatic were drawn from beta distributions with parameters given in Table 1, with means of 2.29% and 17.9%, respectively. We jointly estimated the probability of detection of imported symptomatic infections, ρ_travel_, and the relative offspring number of asymptomatic infections, *α*, by running the importation and branching process models across a range of values of those parameters and calculating the probability of observing the number of reported deaths through March 12; this approach is described in more detail in the parameter calibration section below. The probability of the number of unobserved imported infections being between 0 and 20,000, along with the 152 observed cases and 1 observed death, was calculated using a multinomial distribution; the number of imported infections was then sampled from that distribution. We then smoothed the date of known imported infections with a Gaussian kernel and sampled dates of all imported infections from that distribution. As an alternative scenario, we distributed the timing of imported infections based on the timing of international incidence, with cases in China excluded after February 3, due to a ban on entrance by non-resident foreign nationals who had been to China within the past 14 days enacted on February 2. For each scenario and parameter combination, we generated 1,000 sets of imported infections.

### Transmission model

We simulated local transmission in the United States from January 1 to March 12 using a branching process model, seeded by the aforementioned importation model. Each replicate draw of the number and timing of imported infections seeded one simulation of the branching process model, to maximally represent uncertainty in both importation and transmission processes. The number of secondary infections generated by each infection in the branching process model was drawn from a negative binomial offspring distribution with mean R and dispersion parameter k. Under our baseline scenario, we used a dispersion parameter of k = 1,000, approximating a Poisson distribution, due to a lack of estimates of k for SARS-CoV-2. Under alternative scenarios for k, we considered values of 0.15 and 0.30 to account for superspreading observed in outbreaks of SARS and MERS (33, 34). The number of secondary infections generated by asymptomatic individuals was also drawn from a negative binomial distribution, but with mean *α*R, where *α* in [0,1]. Whether an individual was symptomatic was determined by a Bernoulli trial with probability equal to the proportion of infections that were asymptomatic in that replicate. Each secondary infection’s exposure time was drawn from a log-normal generation interval distribution with mean 4.56 days. In doing so, we assumed that the generation interval followed the same distribution as the serial interval.

In addition to exposure, we simulated three additional outcomes, and the timing thereof, in a subset of infections.

- <u>Symptom onset:</u> The number of new symptomatic infections on day t was drawn from a binomial distribution with the number of trials equal to the number of infections with time of potential symptom onset on day t, and the probability of success equal to the proportion of infections that are symptomatic. For infections that were simulated to result in symptoms, the time of symptom onset was drawn from a Weibull incubation period distribution with mean 7.07 (3) and added to each individual’s exposure time.
- <u>Case reporting:</u> The number of cases reported on day t was drawn from a binomial distribution with the number of trials equal to the number of infections with time of potential case reporting on day t, and the probability of success equal to the proportion of infections that are symptomatic. This accounts for the delay in reporting, but not underreporting, which is addressed below when we calculate the probability that a symptomatic infection is detected, ρ_local_. The time of potential case reporting was drawn from a gamma distribution of the period between symptom onset and case reporting with mean 6 days, and added to each infection’s time of symptom onset.
- <u>Death:</u> The number of deaths on day t was drawn from a binomial distribution with the number of trials equal to the number of infections that could have experienced death on day t, and the probability of success equal to the case fatality risk. The time of death was drawn from a log-normal distribution of time from symptom onset to death with mean 14 days (35), and added to each individual’s time of symptom onset.

All parameter values, and their associated distributions, are described in Table 1. Where parameter distributions were described in the literature using medians and interval measures of spread, we used the optim function in R to estimate parameters of those distributions that matched distribution moments reported by those studies. In that sense, all parameters in our analysis were treated as random variables, with associated uncertainty accounted for throughout our analysis. For the delay between symptom onset and case notification, we fitted a gamma distribution to data on the delay between symptoms and reporting for 26 US cases in the MIDAS line list data; the gamma distribution fitted the data better than negative binomial or log-normal distributions according to AIC (133.5, 134.6, and 134.0, respectively) (Fig. S1). Our mean estimate of 6.0 for this delay is in line with previous estimates from China of 5.8 by Li et al. (36) and 5.5 by Bi et al. (37). Three key parameters – R, the serial interval, and the incubation period – were taken from a single reference (3) to ensure that those estimates were consistent with each other. That is important because R and the serial interval jointly control the epidemic growth rate (38), so taking estimates of R and the serial interval from different studies could have led to unrealistic projections of epidemic growth rate.

We estimated how the probability of detecting locally acquired, symptomatic infections, ρ_local_, changed over time. These estimates were based on the number of symptomatic cases reported each day, C(t), and our model’s predictions for the number of symptomatic infections that could have been reported each day, S(t), after accounting for a delay between symptom onset and reporting. We assumed a uniform prior for ρ_local_, and on each day estimated a posterior equal to ρ_local_(t) ∼ Beta(1+C(t), 1+S(t)-C(t)). We then smoothed over each of 1,000 replicates of independent daily draws of logit-transformed values of ρ_local_(t) using the smooth.spline function in the stats package in R, using weekly knots (Fig. S2).

To understand how many deaths may occur after the time period of our analysis based on infections occurring through then, we set R=0 from March 13 onwards and simulated our model forward to May 31. This allowed any infections occurring by March 12 enough time to result in death, for the proportion expected to result in that outcome.

### Parameter calibration

Due to a lack of prior estimates for two parameters, we jointly estimated the proportion of imported symptomatic infections that were detected, ρ_travel_, and the relative infectiousness of asymptomatic infections, *α*. We fitted these parameters to the total number of deaths resulting from locally acquired SARS-CoV-2 infections in the US by March 12. To approximate a likelihood for given values of ρ_travel_ and *α*, we simulated 200 replicate time series of imported infections, each based on the same value of ρ_travel_, and then simulated local transmission using the same value of *α* for each of the 200 replicates. For each of these 200 replicate simulations, we calculated the cumulative number of infections, I_D_, that, based on their timing, could have resulted in death by March 12. We then calculated the likelihood of the reported number of deaths, D, according to a binomial distribution in which D is the number of successes among I_D_ trials that each have probability of success IFR, where IFR is equal to the CFR times one minus the probability of being asymptomatic. Each of the 200 replicates used independent draws from the uncertainty distributions of other model parameters, so we took the average of the 200 likelihoods to obtain a single marginal likelihood for a given value of ρ_travel_ and *α*. After calculating this marginal likelihood across a grid of values between 0 (or 0.01 for ρ_travel_) and 1 in increments of 0.05 for each parameter, we smoothed this marginal likelihood surface using the bicubic.grid function in the akima package in R (39) to create a gridded marginal likelihood surface with a 0.001 x 0.001 mesh. Finally, we drew samples from the posterior probability distribution of these parameters by resampling from this smoothed marginal likelihood surface, which implicitly assumed a uniform prior on the two parameters. We repeated this calibration procedure for each scenario that we explored, obtaining different estimates for ρ_travel_ and *α* for each of our sensitivity analyses.

### Sensitivity analysis

In addition to the alternative importation models, we also undertook a one-at-a-time sensitivity analysis for each parameter shown in Table 1, with the exception of the calibrated parameters (the last two rows). These last two parameters were re-calibrated as described in the previous section for each new parameter set and importation timing combination. Including the baseline scenario, there were a total of 18 scenarios (i.e., the baseline plus two explored values for each of seven parameters plus one additional scenario with different importation timing). For some parameter values explored in sensitivity analyses, we did not directly use literature estimates, but instead chose values which were plausible minima or maxima for that parameter; these are indicated by “lower” or “higher” in Table 1. For the dispersion parameter, we wanted to explore a value that allowed for superspreading but that generated less overdispersion than was observed for SARS; this formed our intermediate value in the sensitivity analysis. All baseline values were taken directly from literature estimates, with the exception of reporting delay, which was calibrated as described in the branching process model section. For that parameter, we obtained the low and high scenarios by multiplying the shape parameter by 0.5 and 1.5, respectively, while keeping the rate parameter the same. In this way, the reporting delay is the sum of one, two, or three identically distributed gamma random variables in the low, baseline, and high scenarios, respectively.

## Data Availability

All code and data used are available at http://github.com/TAlexPerkins/sarscov2_unobserved.

## Acknowledgments

R.J.O. was supported by an Arthur J. Schmitt Fellowship and an Eck Institute for Global Health Fellowship, and M.P. was supported by a Richard and Peggy Notebaert Premier Fellowship. We thank Jason Rohr and Moritz Kraemer for feedback on the manuscript.

## Supplementary Information Text

### Imported infection predictions and estimates of imported case detection probability

By March 12, there were a total of 152 reported cases and one reported death in the US that were classified as imported on the basis of international travel to areas with known SARS-CoV-2 transmission (19). By jointly estimating ρ_travel_ and the relative infectiousness of asymptomatic infections, *α*, we obtained a median estimate of 0.39 (95% PPI: 0.15 - 0.90) for ρ_travel_ under our baseline scenario. This resulted in a median of 452 (95% PPI: 206 - 1068) imported infections. Under the alternative importation timing scenario, where importation timing was based on international case reports, we obtained a median estimate of 1.00 (95% PPI: 0.98 - 1.00) for ρ_travel_ and 187 (95% PPI: 174 - 202) imported infections. An estimate of ρ_travel_ = 1.00 implies that all symptomatic imported infections were detected, but it still means that asymptomatic infections would have been undetected. Whereas the baseline importation scenario resulted in most importations happening in March and a few throughout February and January, the alternative importation scenario resulted in many importations still happening in March but a large proportion of them also happening around late January (Fig. S3).

### Posterior predictive check against reported local cases

Using our estimate of ρ_local_(t), we simulated the number of reported cases through time and compared this with the actual number of reported cases. By March 12, our model predicted that there should have been 1,546 (95% PPI: 476 - 3,611) reported cases, commensurate with the actual number of 1,514 reported cases (Fig. S4). As expected, this confirms that our estimates of ρ_local_(t) were consistent with the model and the data.

### Sensitivity analysis of unknown parameters

Estimates of the proportion of imported symptomatic infections that were detected, ρ_travel_, and the infectiousness of asymptomatic infections relative to symptomatic infections, *α*, varied based on the values of the other parameters. In general, higher values for parameters expected to increase transmission (e.g., R) were associated with higher estimates of ρ_travel_ (Table S1). Compared to a baseline median estimate of ρ_travel_ = 0.39 (95% PPI: 0.15 - 0.90) with R = 1.97, the estimate of ρ_travel_ was 0.83 (95% PPI: 0.47 - 0.99) with R = 2.7 and 0.08 (95% PPI: 0.04 - 0.19) with R = 1.5. For a shorter serial interval with a mean of 4.7 days, the estimate was ρ_travel_ = 0.52 (95% PPI: 0.19 - 0.96), and with a longer mean serial interval of 7.5 days, the estimate was 0.06 (95% PPI: 0.03 - 0.14). The estimated value of ρ_travel_ was also lower if the CFR was low (ρ_travel_ = 0.20, 95% PPI: 0.08 - 0.53), compared to the scenario with a higher CFR (ρ_travel_ = 0.54, 95% PPI: 0.21 - 0.96). Higher ρ_travel_ estimates correspond to fewer undetected imported infections; therefore, fewer undetected importations are required to account for the observed number of local deaths through March 12 if the CFR is high, R is high, or the serial interval is short. In addition, when we based the timing of importations on international incidence (excluding China after travel restrictions were implemented on February 3) the estimate of ρ_travel_ was 1.00 (95% PPI: 0.98 - 1.00) due to the increased probability of early importations – and more time for local infections to increase – under this scenario. There was greater uncertainty in our *α* estimates under most sensitivity scenarios, and in most scenarios the estimates of ρ_travel_ and *α* were positively correlated (Fig. S5).

### Sensitivity analysis of cumulative infections

Because ρ_travel_ and *α* were estimated for each parameter-sensitivity scenario, cumulative infections were relatively similar under the low, baseline, and high scenarios for many parameters. Cumulative infections were most sensitive to assumptions about R, the serial interval, and the timing of imported infections (Fig. S6, Table S2). The former two affect how quickly local infections increase, and the latter affects how much time they have to increase. Cumulative infections were also somewhat sensitive to assumptions about case fatality risk and the delay between exposure and death, because assumptions about those parameters influenced estimates of ρ_travel_ and *α*, which were based on reported deaths.

### Sensitivity analysis of local case detection probability

The proportion of symptomatic infections detected over time followed a similar pattern under all parameter sensitivity scenarios, with low values of ρ_local_ throughout late February followed by increases in March (Fig. S7). Long delays in case detection (9 days) were associated with the lowest proportion of symptomatic infections detected; in that scenario, ρ_local_ mostly did not exceed 10%.

### Sensitivity analysis of the ratio of deaths after and before March 12

The ratio of deaths expected March 13 and after, relative to before then, was higher with changes in parameters that resulted in faster growth in local infections and later arrival of imported infections (Fig. S8, Table S3). The proportion of deaths expected to occur after March 12 also increased with increases in the delay between symptom onset and death (Table S3). Overdispersion (lower k) did not drastically alter our estimates of ρ_travel_ or *α* (Table S1) or the number of cumulative infections (Table S2), but it did extend the lower and upper bounds on the range of the ratio of deaths after and before March 12 (Table S3).

**Figure S1.**
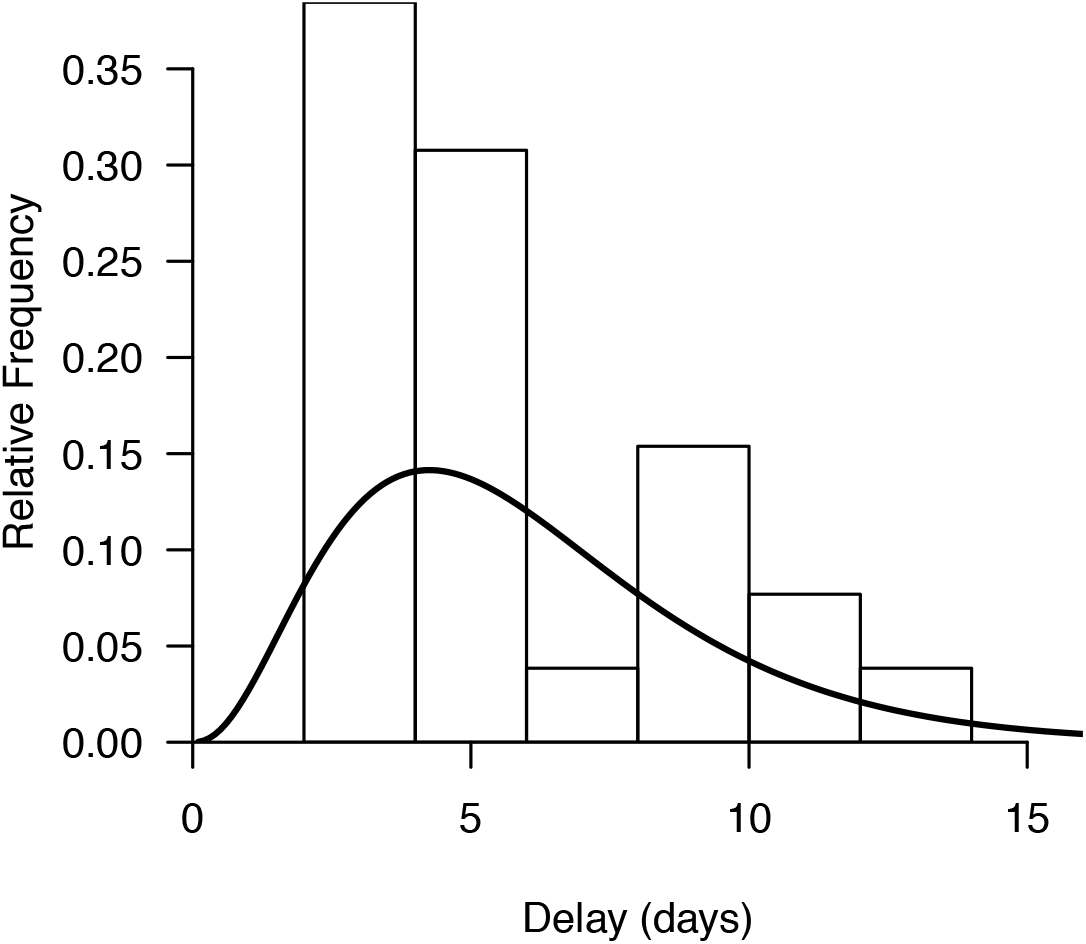
Distribution of the delay between symptom onset and reporting for 26 US cases. The curve shows the maximum-likelihood fit of a gamma distribution (shape = 3.43, rate = 0.572) to those data.

**Figure S2.**
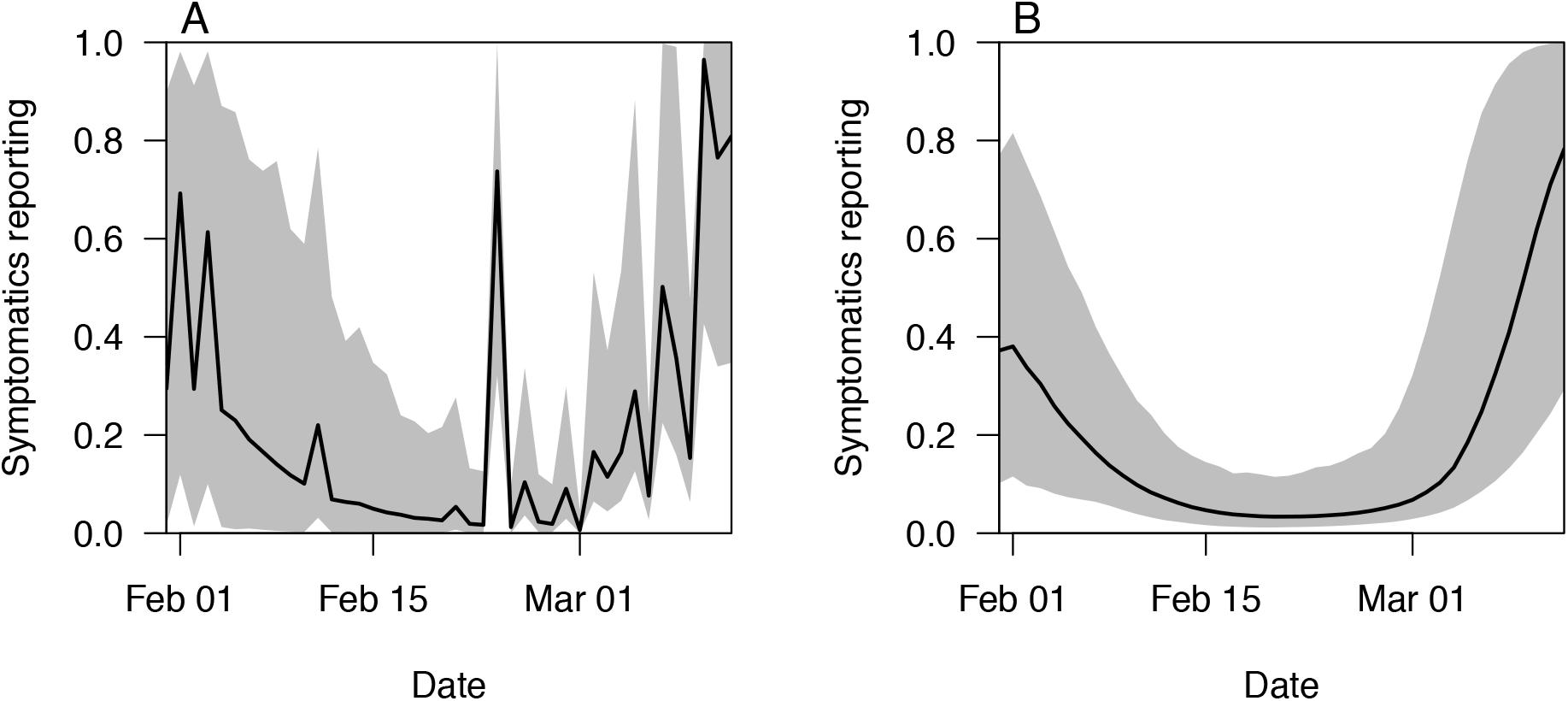
Comparison of daily estimates of ρ_local_(*t*) with and without smoothing. The two panels compare A) raw estimates with no smoothing and B) smoothed estimates with splines. We used the smoothed estimates in our analysis given that they are more indicative of general trends in case detection.

**Figure S3.**
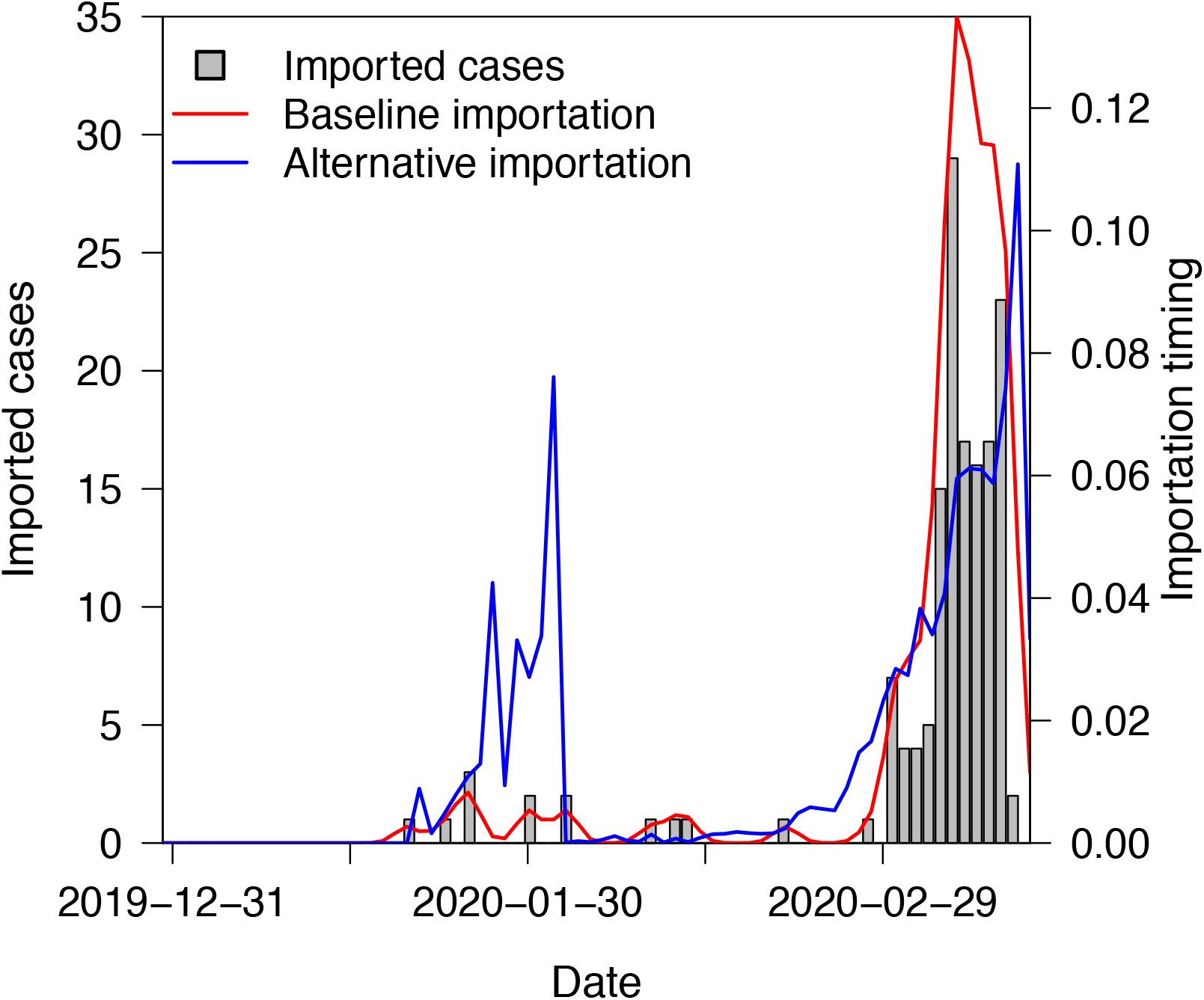
Assumptions about timing of imported infections. Imported cases that have been reported are shown in gray, and the red line shows the baseline distribution of timing of imported infections that we based on a Gaussian kernel smooth of those data. The blue line shows an alternative distribution of timing of imported infections based on patterns of international incidence.

**Figure S4.**
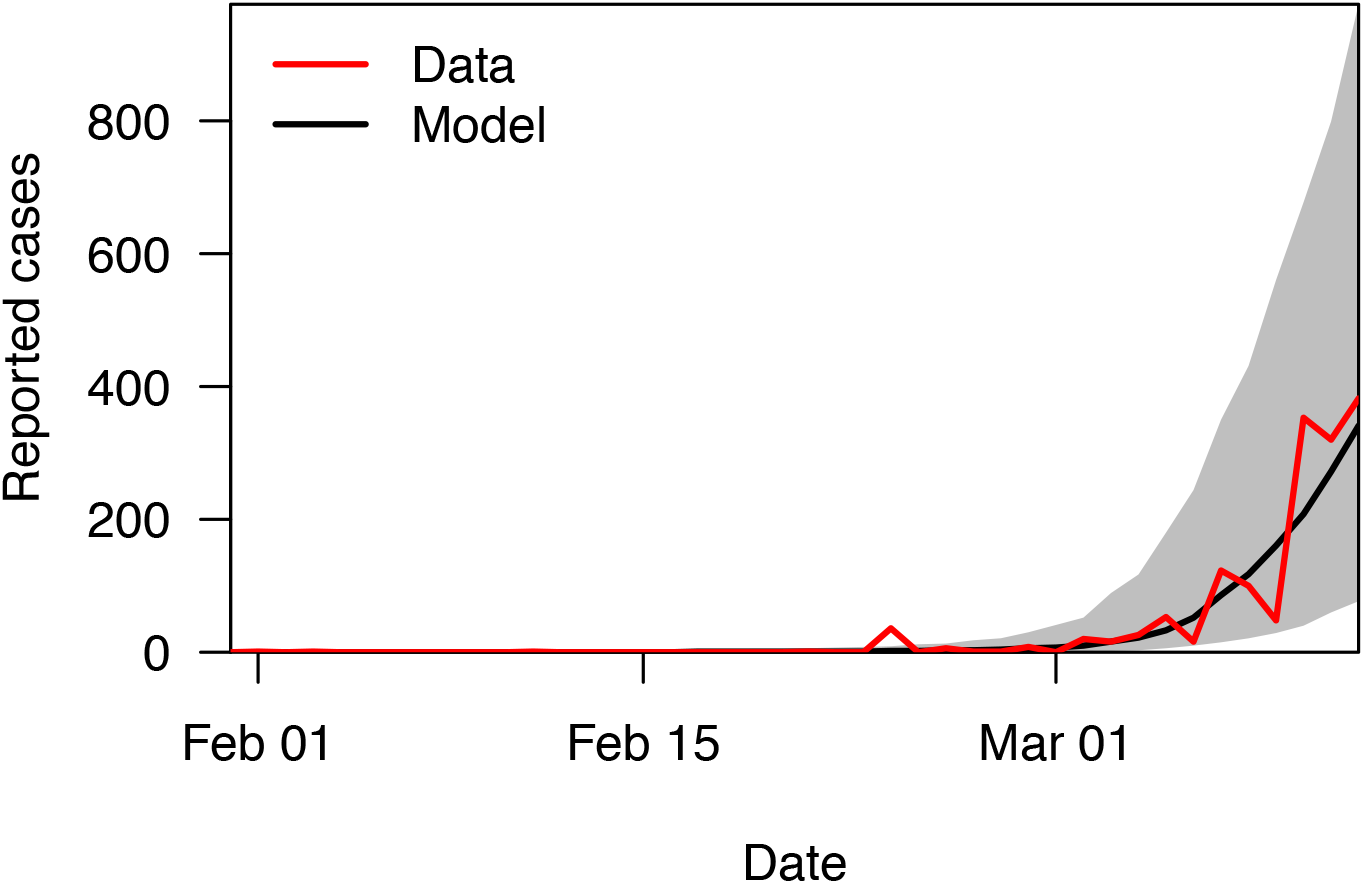
The number of cases reported in the US compared to the number our model predicts were reported.

**Figure S5.**
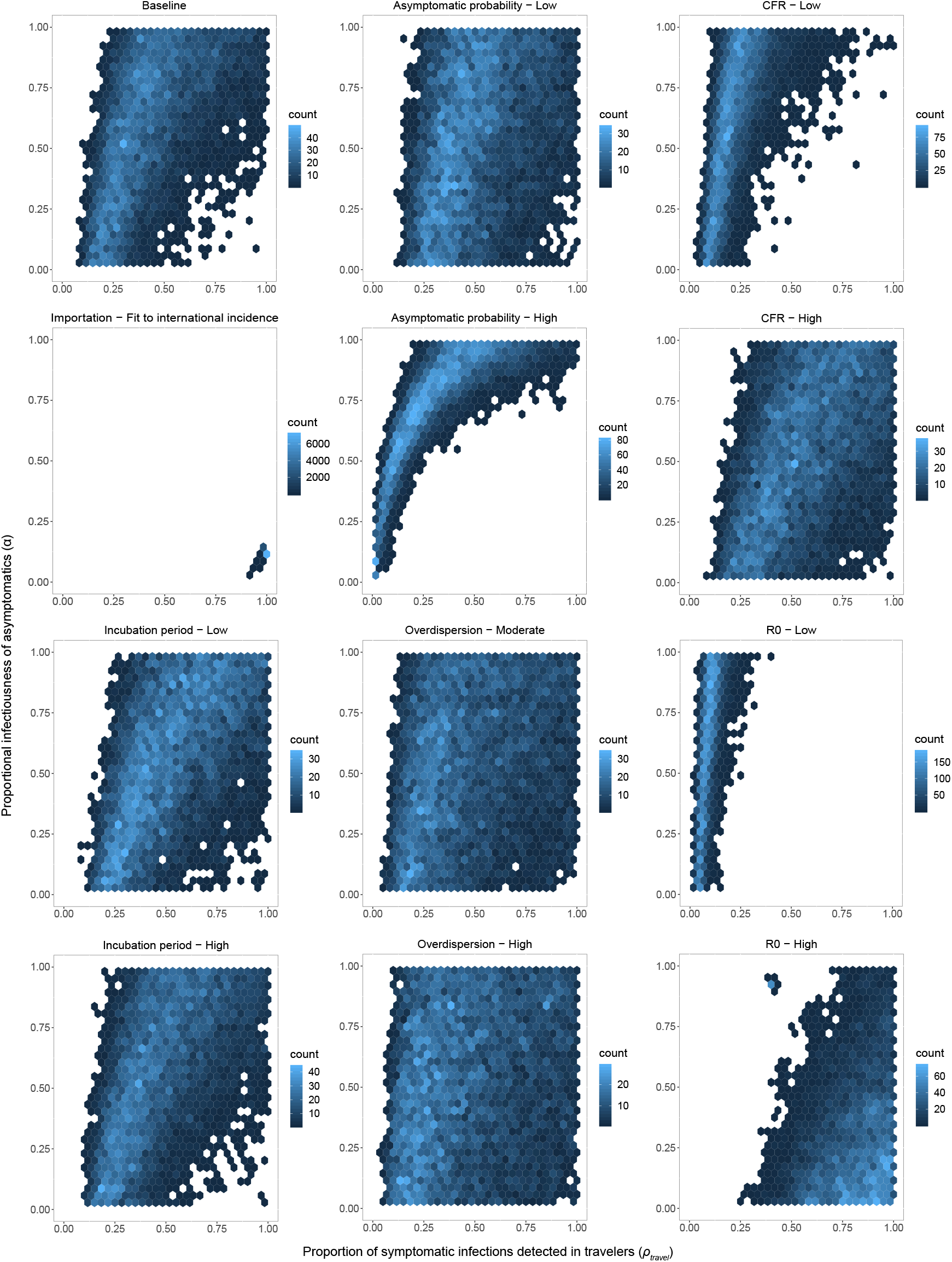
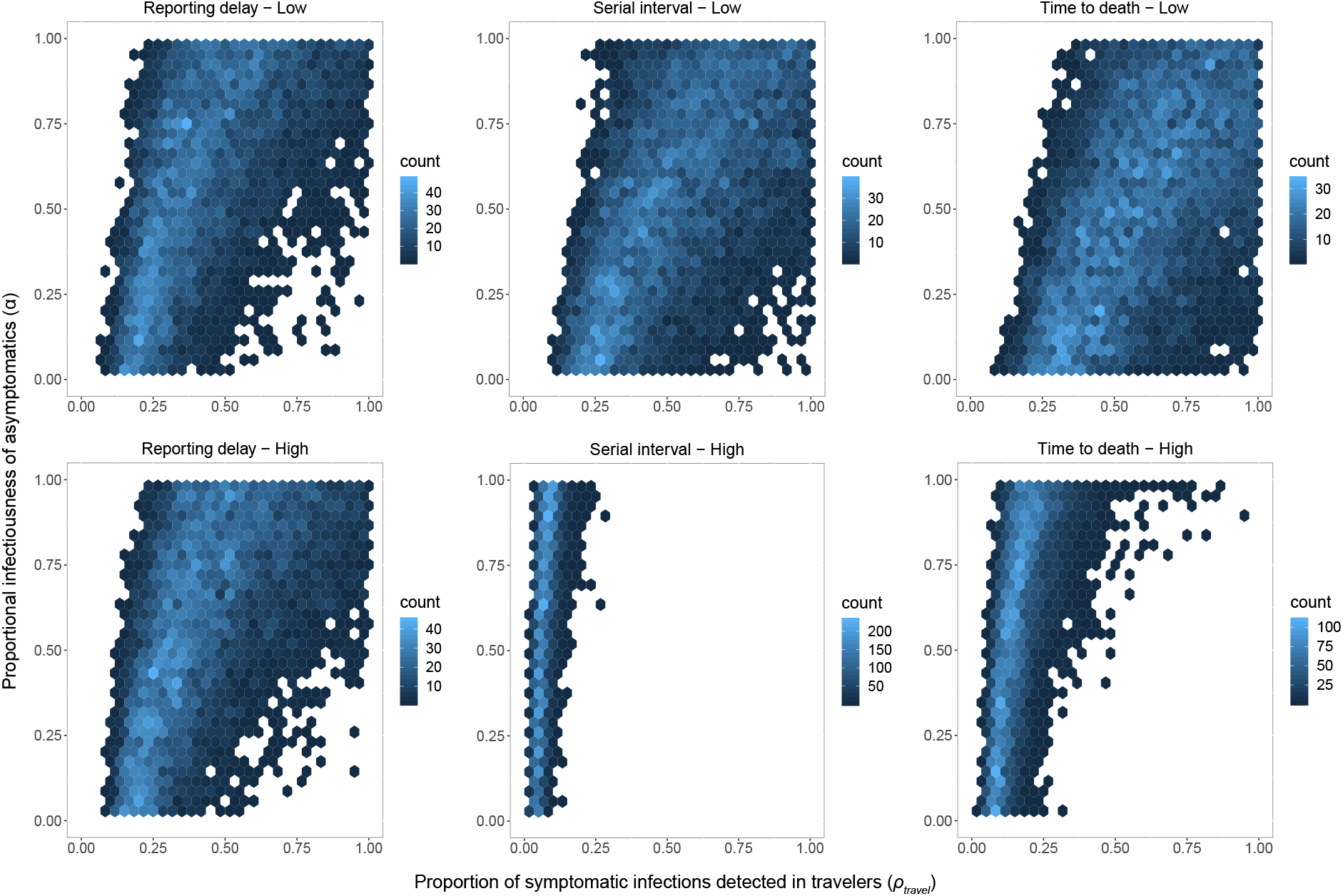
Samples (10^4^) from the joint posterior distribution of the proportion of imported symptomatic infections detected (ρ_travel_) and the relative infectiousness of asymptomatic infections (α) under different parameter-sensitivity scenarios.

**Figure S6.**
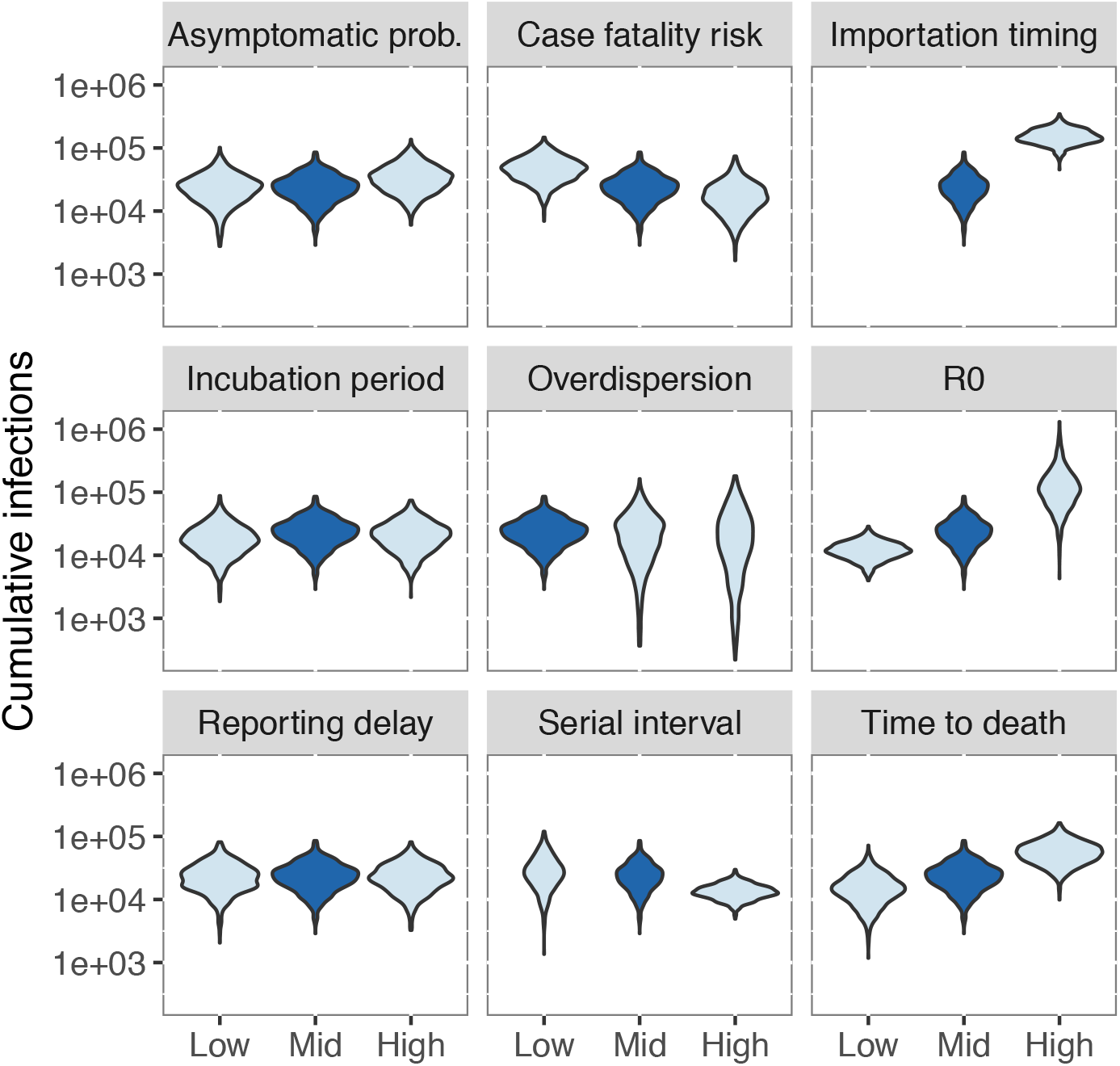
Posterior predictive distributions of cumulative infections by March 12 under different parameter sensitivity scenarios. Unlike other parameters, importation timing was not described in terms of simple numerical values; in that case, “mid” refers to our baseline assumption that the timing of unobserved imported infections followed the timing of observed imported cases, and “high” refers to the alternative scenario that their timing followed international incidence patterns.

**Figure S7.**
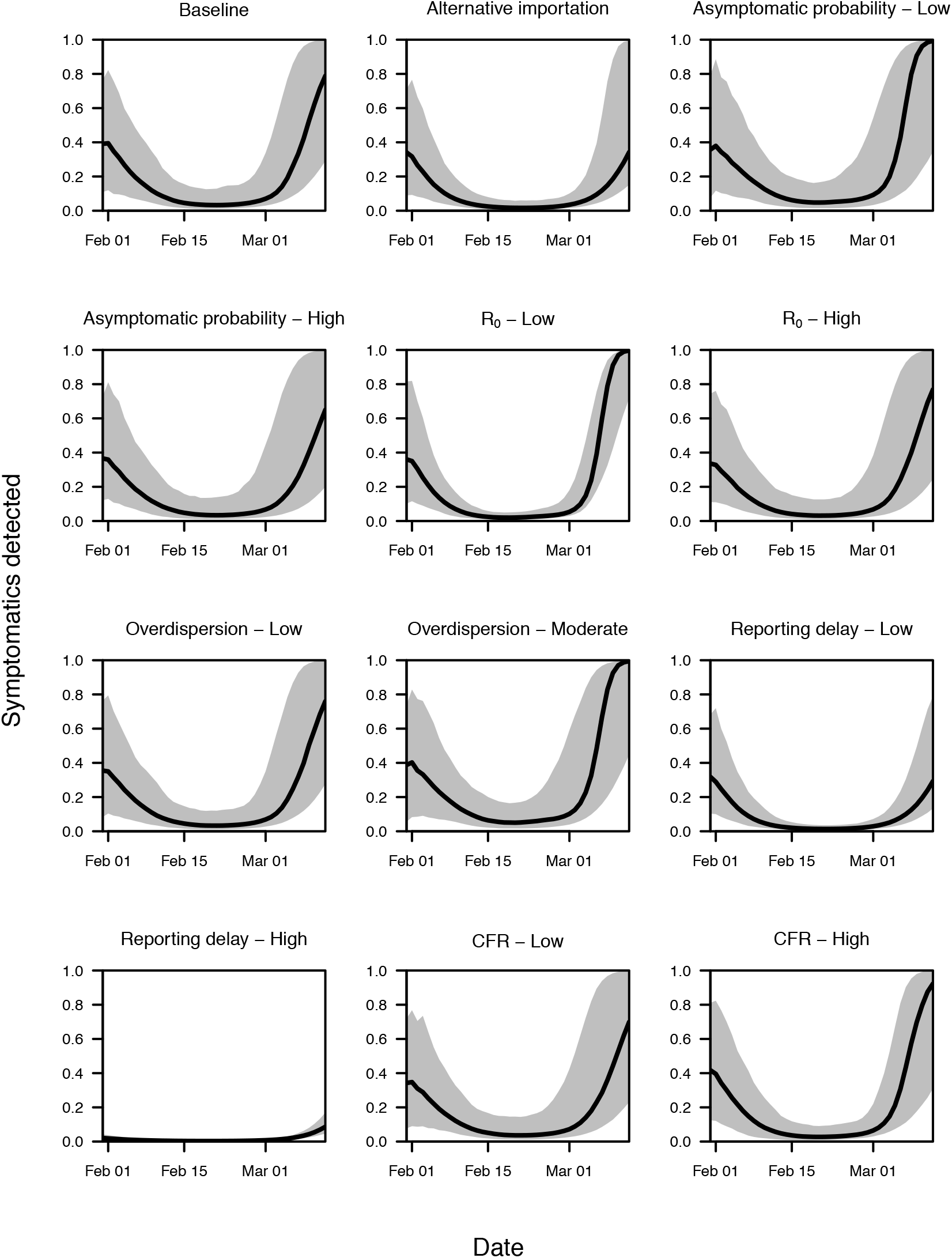
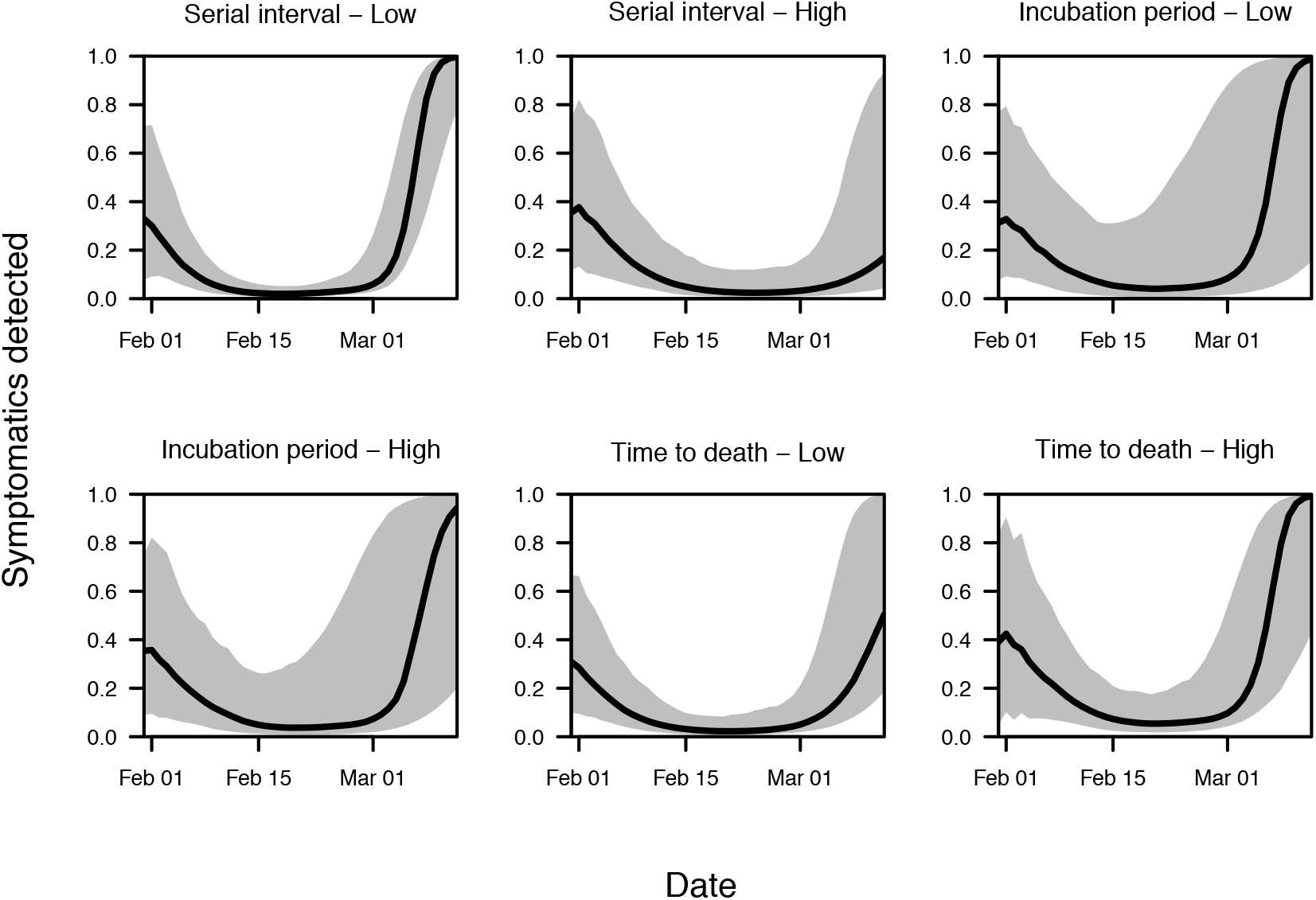
Median and 95% posterior predictive interval of the probability of detecting a local symptomatic infection, ρ_local_(*t*), after accounting for delays in reporting. Each panel represents a different parameter-sensitivity scenario.

**Figure S8.**
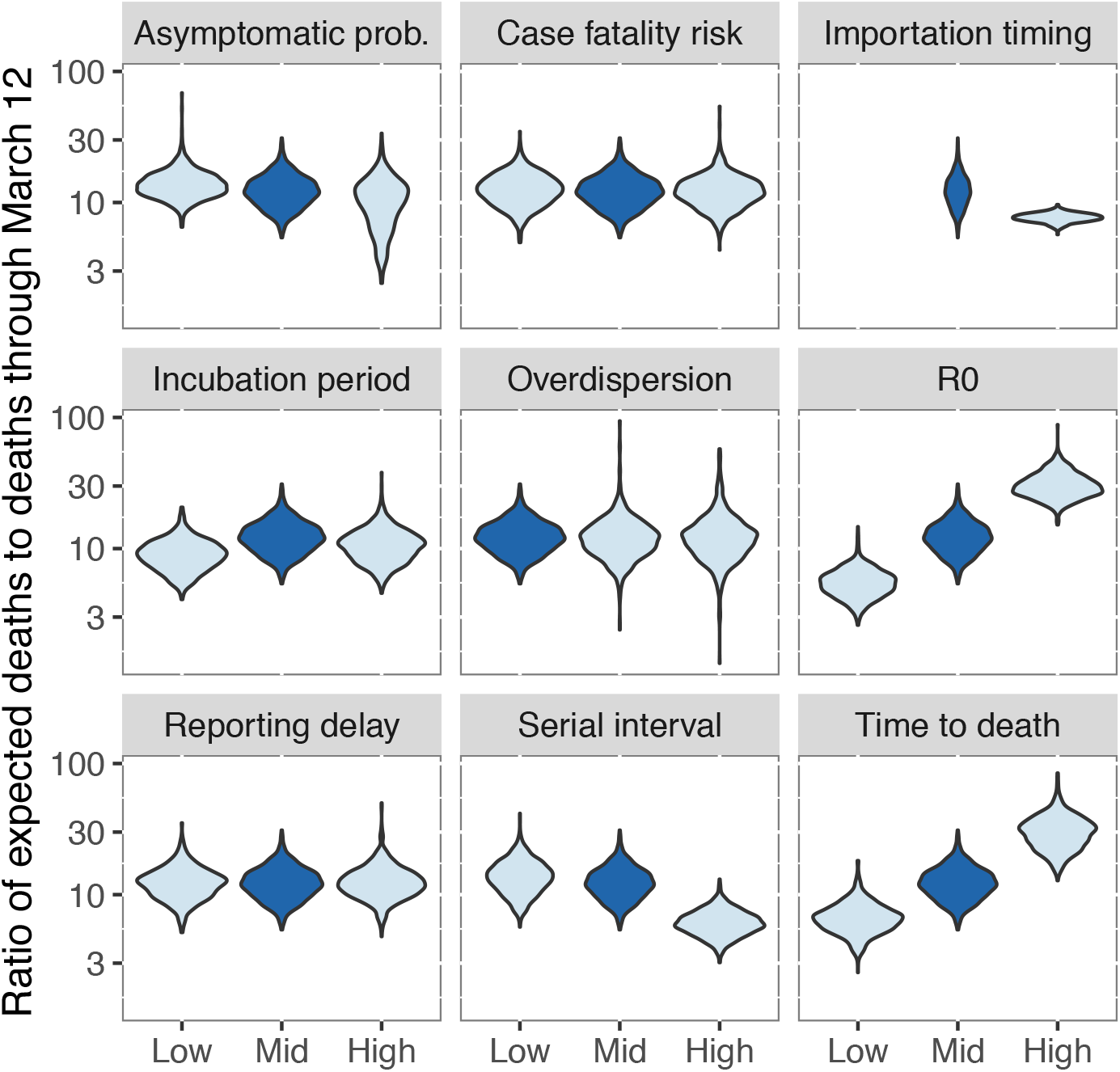
Posterior predictive distributions of the ratio of deaths after and before March 12 under different parameter sensitivity scenarios. Unlike other parameters, importation timing was not described in terms of simple numerical values; in that case, “mid” refers to our baseline assumption that the timing of unobserved imported infections followed the timing of observed imported cases, and “high” refers to the alternative scenario that their timing followed international incidence patterns.

**Table S1.** Median estimates and 95% posterior predictive intervals of the marginal distributions of proportion of imported symptomatic infections detected (ρ_travel_) and the relative infectiousness of asymptomatic infections (α) under different parameter-sensitivity scenarios.

| Parameter varied | Scenario | Prob. symptomatic infections detected ($\rho_{\text{travel}}$) | | | Relative asymptomatic infectiousness ($\alpha$) | | |
| --- | --- | --- | --- | --- | --- | --- | --- |
| Baseline |  | 0.39 | (0.15 | - 0.90) | 0.61 | (0.04 | - 0.98) |
| Asymptomatic prop. | Low | 0.47 | (0.21 | - 0.93) | 0.55 | (0.03 | - 0.98) |
| Asymptomatic prop. | High | 0.29 | (0.01 | - 0.81) | 0.79 | (0.10 | - 0.99) |
| R | Low | 0.08 | (0.04 | - 0.19) | 0.63 | (0.04 | - 0.99) |
| R | High | 0.83 | (0.47 | - 0.99) | 0.23 | (0.01 | - 0.89) |
| Dispersion, k | Moderate | 0.41 | (0.13 | - 0.96) | 0.54 | (0.03 | - 0.98) |
| Dispersion, k | High | 0.41 | (0.11 | - 0.95) | 0.53 | (0.03 | - 0.98) |
| Reporting delay | Low | 0.39 | (0.15 | - 0.90) | 0.60 | (0.04 | - 0.98) |
| Reporting delay | High | 0.39 | (0.15 | - 0.90) | 0.61 | (0.04 | - 0.98) |
| CFR | Low | 0.20 | (0.08 | - 0.53) | 0.64 | (0.05 | - 0.99) |
| CFR | High | 0.54 | (0.21 | - 0.96) | 0.54 | (0.04 | - 0.98) |
| Serial interval | Low | 0.52 | (0.19 | - 0.96) | 0.54 | (0.04 | - 0.97) |
| Serial interval | High | 0.06 | (0.03 | - 0.14) | 0.61 | (0.04 | - 0.98) |
| Incubation period | Low | 0.50 | (0.20 | - 0.96) | 0.57 | (0.04 | - 0.98) |
| Incubation period | High | 0.44 | (0.17 | - 0.93) | 0.58 | (0.03 | - 0.98) |
| Time to death | Low | 0.58 | (0.23 | - 0.97) | 0.51 | (0.03 | - 0.97) |
| Time to death | High | 0.15 | (0.06 | - 0.36) | 0.61 | (0.04 | - 0.98) |
| Importation timing | High | 1.00 | (0.98 | - 1.00) | 0.13 | (0.11 | - 0.14) |

**Table S2.**
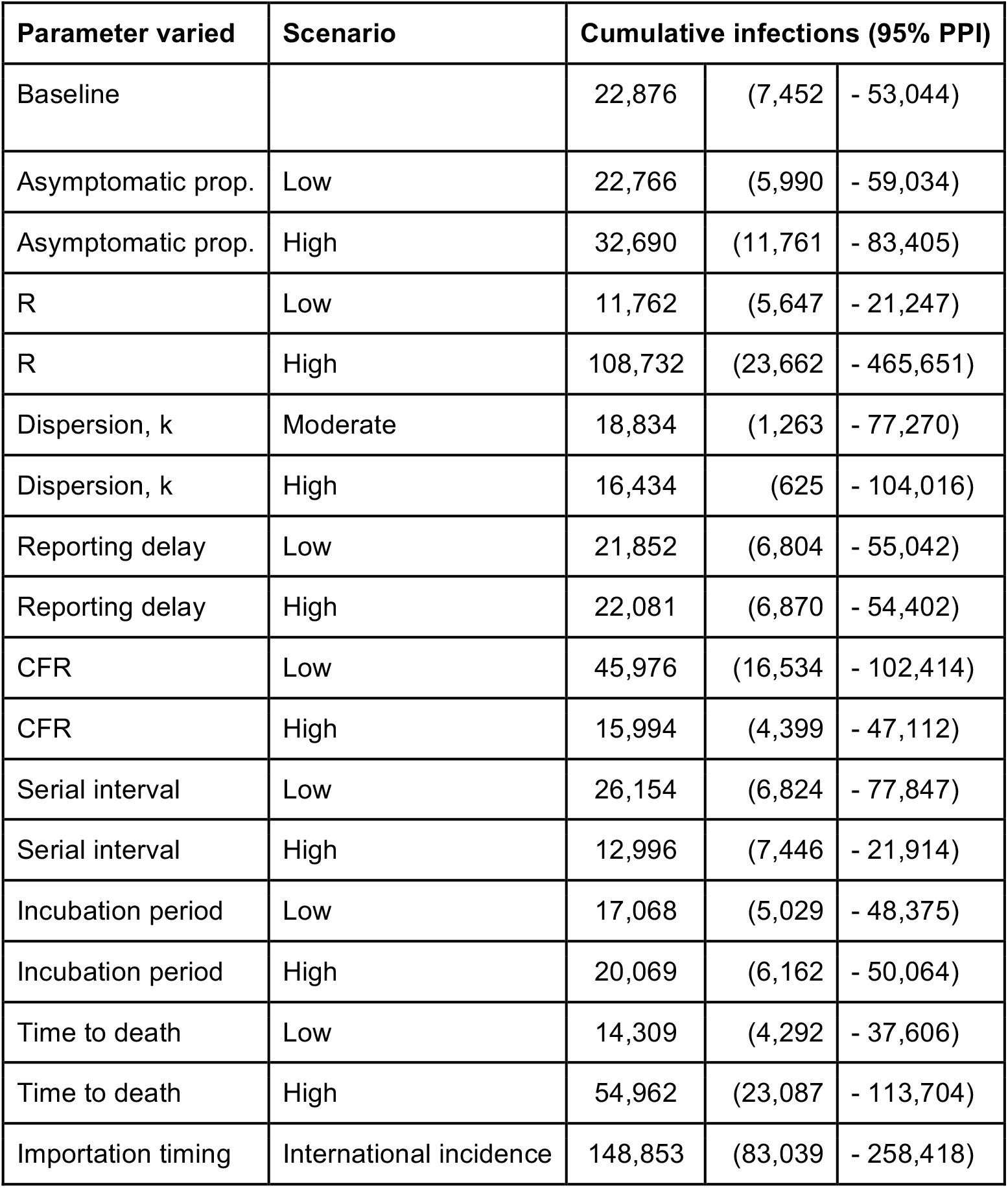
Median estimates and 95% posterior predictive intervals of cumulative infections under different parameter sensitivity scenarios.

| Parameter varied | Scenario | Cumulative infections (95% PPI) |  |  |
| --- | --- | --- | --- | --- |
| Baseline |  | 22,876 | (7,452 | - 53,044) |
| Asymptomatic prop. | Low | 22,766 | (5,990 | - 59,034) |
| Asymptomatic prop. | High | 32,690 | (11,761 | - 83,405) |
| R | Low | 11,762 | (5,647 | - 21,247) |
| R | High | 108,732 | (23,662 | - 465,651) |
| Dispersion, k | Moderate | 18,834 | (1,263 | - 77,270) |
| Dispersion, k | High | 16,434 | (625 | - 104,016) |
| Reporting delay | Low | 21,852 | (6,804 | - 55,042) |
| Reporting delay | High | 22,081 | (6,870 | - 54,402) |
| CFR | Low | 45,976 | (16,534 | - 102,414) |
| CFR | High | 15,994 | (4,399 | - 47,112) |
| Serial interval | Low | 26,154 | (6,824 | - 77,847) |
| Serial interval | High | 12,996 | (7,446 | - 21,914) |
| Incubation period | Low | 17,068 | (5,029 | - 48,375) |
| Incubation period | High | 20,069 | (6,162 | - 50,064) |
| Time to death | Low | 14,309 | (4,292 | - 37,606) |
| Time to death | High | 54,962 | (23,087 | - 113,704) |
| Importation timing | International incidence | 148,853 | (83,039 | - 258,418) |

**Table S3.**
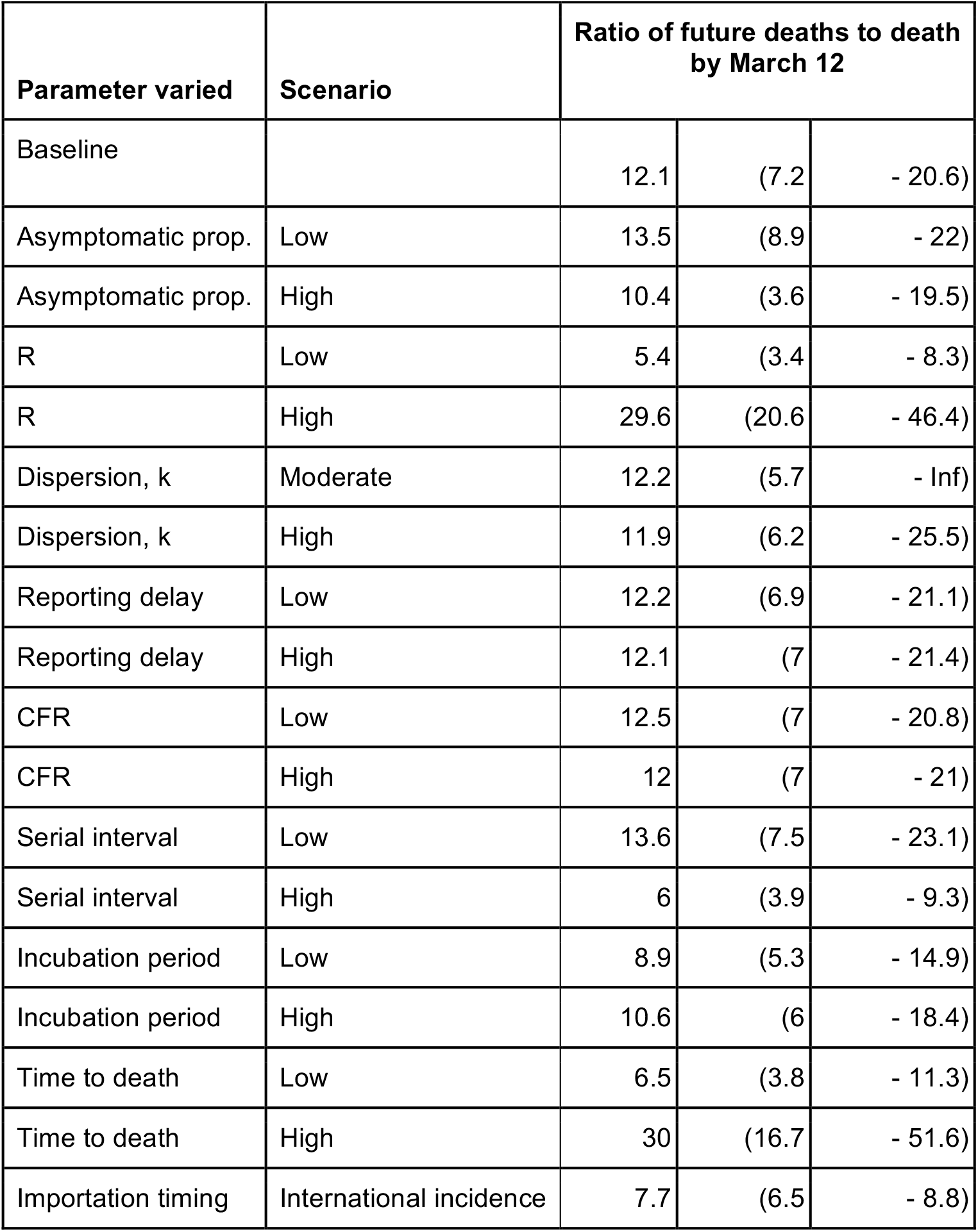
Median estimates and 95% posterior predictive intervals of the ratio of deaths after and before March 12 under different parameter sensitivity scenarios.

## Notes

### Competing Interest Statement

The authors have declared no competing interest.

### Funding Statement

RJO acknowledges support from an Arthur J. Schmitt Leadership Fellowship in Science and Engineering and an Eck Institute for Global Health Fellowship.
MP acknowledges support from a Richard and Peggy Notebaert Premier Fellowship.

## References

1. World Health Organization, “Coronavirus disease 2019 (COVID-19) Situation Report” (51, 2020).

2. N. van Doremalen, T. Bushmaker, D. Morris, M. Holbrook, A. Gamble, B. Williamson, A. Tamin, J. Harcourt, N. Thornburg, S. Gerber, J. Lloyd-Smith, E. de Wit, V. Munster, Aerosol and surface stability of HCoV-19 (SARS-CoV-2) compared to SARS-CoV-1. medRxiv, 2020.03.09.20033217 (2020).

3. L. Tindale, M. Coombe, J. E. Stockdale, E. Garlock, W. Y. V. Lau, M. Saraswat, Y.-H. B. Lee, L. Zhang, D. Chen, J. Wallinga, Others, Transmission interval estimates suggest pre-symptomatic spread of COVID-19. medRxiv (2020).

4. Y. Bai, L. Yao, T. Wei, F. Tian, D.-Y. Jin, L. Chen, M. Wang, Presumed Asymptomatic Carrier Transmission of COVID-19. JAMA (2020).

5. K. Mizumoto, K. Kagaya, A. Zarebski, G. Chowell, Estimating the Asymptomatic Proportion of 2019 Novel Coronavirus onboard the Princess Cruises Ship, 2020. medRxiv (2020).

6. Z. Wu, J. M. McGoogan, Characteristics of and Important Lessons From the Coronavirus Disease 2019 (COVID-19) Outbreak in China: Summary of a Report of 72 314 Cases From the Chinese Center for Disease Control and Prevention. JAMA (2020).

7. R. Li, S. Pei, B. Chen, Y. Song, T. Zhang, W. Yang, J. Shaman, Substantial undocumented infection facilitates the rapid dissemination of novel coronavirus (SARS-CoV2). Science (2020).

8. M. Gilbert, G. Pullano, F. Pinotti, E. Valdano, C. Poletto, P.-Y. Boëlle, E. D’Ortenzio, Y. Yazdanpanah, S. P. Eholie, M. Altmann, B. Gutierrez, M. U. G. Kraemer, V. Colizza, Preparedness and vulnerability of African countries against importations of COVID-19: a modelling study. Lancet (2020).

9. CDC, Coronavirus Disease 2019 (COVID-19). Centers for Disease Control and Prevention (2020), (available at https://web.archive.org/web/20200227172220/ http://www.cdc.gov/coronavirus/2019-nCoV/hcp/clinical-criteria.html).

10. R. M. Burke, Active Monitoring of Persons Exposed to Patients with Confirmed COVID-19 — United States, January–February 2020. MMWR Morb. Mortal. Wkly. Rep. 69 (2020).

11. A. Maxmen, How much is coronavirus spreading under the radar? Nature (2020).

12. Y. Ng, Evaluation of the Effectiveness of Surveillance and Containment Measures for the First 100 Patients with COVID-19 in Singapore — January 2–February 29, 2020. MMWR Morb. Mortal. Wkly. Rep. 69 (2020).

13. Division of Risk assessment and International cooperation, 76 additional cases have been confirmed. The Updates of COVID-19 in Republic of Korea, (available at https://www.cdc.go.kr/board/board.es?mid=&bid=0030#).

14. Proclamation on Declaring a National Emergency Concerning the Novel Coronavirus Disease (COVID-19) Outbreak |The White House. The White House, (available at https://www.whitehouse.gov/presidential-actions/proclamation-declaring-national-emergency-concerning-novel-coronavirus-disease-covid-19-outbreak/).

15. The Associated Press, Americans Brace for New Life of No School and Growing Dread. The New York Times (2020), (available at https://www.nytimes.com/aponline/2020/03/14/us/ap-us-virus-outbreak-us.html).

16. J. Hellewell, S. Abbott, A. Gimma, N. I. Bosse, C. I. Jarvis, T. W. Russell, J. D. Munday, A. J. Kucharski, W. J. Edmunds, Feasibility of controlling COVID-19 outbreaks by isolation of cases and contacts. Lancet Global Health (2020).

17. R. M. Anderson, H. Heesterbeek, D. Klinkenberg, T. D. Hollingsworth, How will country-based mitigation measures influence the course of the COVID-19 epidemic? Lancet (2020).

18. E. Dong, H. Du, L. Gardner, An interactive web-based dashboard to track COVID-19 in real time. Lancet Infect. Dis. (2020).

19. midas-network, midas-network/COVID-19. GitHub, (available at https://github.com/midas-network/COVID-19).

20. T. Team, Vital Surveillances: The Epidemiological Characteristics of an Outbreak of 2019 Novel Coronavirus Diseases (COVID-19)—China, 2020. China CDC Weekly. 2, 113–122 (2020).

21. W. Wang, J. Tang, F. Wei, Updated understanding of the outbreak of 2019 novel coronavirus (2019-nCoV) in Wuhan, China. J. Med. Virol. 92, 441–447 (2020).

22. F. Zhou, T. Yu, R. Du, G. Fan, Y. Liu, Z. Liu, J. Xiang, Y. Wang, B. Song, X. Gu, L. Guan, Y. Wei, H. Li, X. Wu, J. Xu, S. Tu, Y. Zhang, H. Chen, B. Cao, Clinical course and risk factors for mortality of adult inpatients with COVID-19 in Wuhan, China: a retrospective cohort study. Lancet. (2020).

23. F. Amanat, T. Nguyen, V. Chromikova, S. Strohmeier, D. Stadlbauer, A. Javier, K. Jiang, G. Asthagiri-Arunkumar, J. Polanco, M. Bermudez-Gonzalez, D. Caplivski, A. Cheng, K. Kedzierska, O. Vapalahti, J. Hepojoki, V. Simon, F. Krammer, A serological assay to detect SARS-CoV-2 seroconversion in humans. medRxiv (2020).

24. A. J. Kucharski, T. W. Russell, C. Diamond, Y. Liu, J. Edmunds, S. Funk, R. M. Eggo, F. Sun, M. Jit, J. D. Munday, N. Davies, A. Gimma, K. van Zandvoort, H. Gibbs, J. Hellewell, C. I. Jarvis, S. Clifford, B. J. Quilty, N. I. Bosse, S. Abbott, P. Klepac, S. Flasche, Early dynamics of transmission and control of COVID-19: a mathematical modelling study. Lancet Infect. Dis. (2020).

25. M. J. Ferrari, S. Bansal, L. A. Meyers, O. N. Bjørnstad, Network frailty and the geometry of herd immunity. Proc. Biol. Sci. 273, 2743–2748 (2006).

26. M. D. Van Kerkhove, A. I. Bento, H. L. Mills, N. M. Ferguson, C. A. Donnelly, A review of epidemiological parameters from Ebola outbreaks to inform early public health decision-making. Sci Data. 2, 150019 (2015).

27. T. A. Perkins, A. S. Siraj, C. W. Ruktanonchai, M. U. G. Kraemer, A. J. Tatem, Model-based projections of Zika virus infections in childbearing women in the Americas. Nat Microbiol. 1, 16126 (2016).

28. M. Chinazzi, J. T. Davis, M. Ajelli, C. Gioannini, M. Litvinova, S. Merler, A. Pastore Y Piontti, K. Mu, L. Rossi, K. Sun, C. Viboud, X. Xiong, H. Yu, M. E. Halloran, I. M. Longini Jr, A. Vespignani, The effect of travel restrictions on the spread of the 2019 novel coronavirus (COVID-19) outbreak. Science (2020).

29. C. Fraser, S. Riley, R. M. Anderson, N. M. Ferguson, Factors that make an infectious disease outbreak controllable. Proc. Natl. Acad. Sci. U. S. A. 101, 6146–6151 (2004).

30. H. Tian, Y. Liu, Y. Li, C.-H. Wu, B. Chen, M. U. G. Kraemer, B. Li, J. Cai, B. Xu, Q. Yang, B. Wang, P. Yang, Y. Cui, Y. Song, P. Zheng, Q. Wang, O. N. Bjornstad, R. Yang, B. Grenfell, O. Pybus, C. Dye, The impact of transmission control measures during the first 50 days of the COVID-19 epidemic in China. medRxiv (2020).

31. B. Cowling, S. Ali, T. Ng, T. Tsang, J. Li, M. Fong, Q. Liao, M. Kwan, S. Lee, S. Chiu, J. Wu, P. Wu, G. Leung, Impact assessment of non-pharmaceutical interventions against COVID-19 and influenza in Hong Kong: an observational study. medRxiv (2020).

32. D. B. Jernigan, CDC COVID-19 Response Team, Update: Public Health Response to the Coronavirus Disease 2019 Outbreak - United States, February 24, 2020. MMWR Morb. Mortal. Wkly. Rep. 69, 216–219 (2020).

33. A. J. Kucharski, C. L. Althaus, The role of superspreading in Middle East respiratory syndrome coronavirus (MERS-CoV) transmission. Eurosurveillance. 20 (2015).

34. J. O. Lloyd-Smith, S. J. Schreiber, P. E. Kopp, W. M. Getz, Superspreading and the effect of individual variation on disease emergence. Nature. 438, 355–359 (2005).

35. W.-J. Guan, Z.-Y. Ni, Y. Hu, W.-H. Liang, C.-Q. Ou, J.-X. He, L. Liu, H. Shan, C.-L. Lei, D. S. C. Hui, B. Du, L.-J. Li, G. Zeng, K.-Y. Yuen, R.-C. Chen, C.-L. Tang, T. Wang, P.-Y. Chen, J. Xiang, S.-Y. Li, J.-L. Wang, Z.-J. Liang, Y.-X. Peng, L. Wei, Y. Liu, Y.-H. Hu, P. Peng, J.-M. Wang, J.-Y. Liu, Z. Chen, G. Li, Z.-J. Zheng, S.-Q. Qiu, J. Luo, C.-J. Ye, S.-Y. Zhu, N.-S. Zhong, Clinical Characteristics of Coronavirus Disease 2019 in China. N. Engl. J. Med. (2020).

36. Q. Li, X. Guan, P. Wu, X. Wang, L. Zhou, Y. Tong, R. Ren, K. S. M. Leung, E. H. Y. Lau, J. Y. Wong, X. Xing, N. Xiang, Y. Wu, C. Li, Q. Chen, D. Li, T. Liu, J. Zhao, M. Liu, W. Tu, C. Chen, L. Jin, R. Yang, Q. Wang, S. Zhou, R. Wang, H. Liu, Y. Luo, Y. Liu, G. Shao, H. Li, Z. Tao, Y. Yang, Z. Deng, B. Liu, Z. Ma, Y. Zhang, G. Shi, T. T. Y. Lam, J. T. Wu, G. F. Gao, B. J. Cowling, B. Yang, G. M. Leung, Z. Feng, Early Transmission Dynamics in Wuhan, China, of Novel Coronavirus–Infected Pneumonia. N. Engl. J. Med. (2020).

37. Q. Bi, Y. Wu, S. Mei, C. Ye, X. Zou, Z. Zhang, X. Liu, L. Wei, S. A. Truelove, T. Zhang, W. Gao, C. Cheng, X. Tang, X. Wu, Y. Wu, B. Sun, S. Huang, Y. Sun, J. Zhang, T. Ma, J. Lessler, T. Feng, Epidemiology and Transmission of COVID-19 in Shenzhen China: Analysis of 391 cases and 1,286 of their close contacts. medRxiv (2020).

38. J. Wallinga, M. Lipsitch, How generation intervals shape the relationship between growth rates and reproductive numbers. Proc. Biol. Sci. 274, 599–604 (2007).

39. H. Akima, A. Gebhardt, akima: Interpolation of Irregularly and Regularly Spaced Data. R package version 0.6-2 (2016).

40. J. T. Wu, K. Leung, G. M. Leung, Nowcasting and forecasting the potential domestic and international spread of the 2019-nCoV outbreak originating in Wuhan, China: a modelling study. Lancet. 395 (2020), pp. 689–697.

41. COVID-19 reports |Faculty of Medicine |Imperial College London, (available at https://www.imperial.ac.uk/mrc-global-infectious-disease-analysis/news--wuhan-coronavirus/).

42. WHO Director-General’s opening remarks at the media briefing on COVID-19 - 3 March 2020, (available at https://www.who.int/dg/speeches/detail/who-director-general-s-opening-remarks-at-the-media-briefing-on-covid-193-march-2020).

43. H. Nishiura, N. M. Linton, A. R. Akhmetzhanov, Serial interval of novel coronavirus (2019- nCoV) infections. medRxiv (2020).

44. S. A. Lauer, K. H. Grantz, Q. Bi, F. K. Jones, Q. Zheng, H. R. Meredith, A. S. Azman, N. G. Reich, J. Lessler, The Incubation Period of Coronavirus Disease 2019 (COVID-19) From Publicly Reported Confirmed Cases: Estimation and Application. Ann. Intern. Med. (2020).

45. Website, (available at https://www.imperial.ac.uk/media/imperial-college/medicine/sph/ide/gida-fellowships/Imperial-College-2019-nCoV-severity-10-02-2020.pdf).

